# An Assessment of the Effects of Coronavirus Disease 2019 (COVID-19) on Community Pharmacy Service Delivery in Lagos State

**DOI:** 10.1101/2024.11.02.24316307

**Authors:** Olufunke G. Ajibola, Arinola Joda

## Abstract

**Background:** Community pharmacists play an important role as frontline health care workers during the COVID-19 pandemic. Therefore, the delivery of pharmaceutical care and the availability of drugs is compromised. The objective of this study was to evaluate the effect of coronavirus disease 2019 (COVID-19) on community pharmacy service delivery in Lagos state.

**Methods:** A cross-sectional study with an online questionnaire created using Google Forms software was sent to community pharmacists in Lagos State, Nigeria via the Association of Community Pharmacists in Nigeria WhatsApp platforms. The sample size was 120 respondents conveniently chosen. Ethical approval was obtained from Health Research Ethics Committee, Lagos University Teaching Hospital (LUTH), Idi-Araba, Lagos State. Collected data was analyzed using Microsoft Excel.

**Results:** A total of 125 respondents participated in the study. The result showed 56% were female respondents and the mean age of respondents was 32.4 ± 4.7 years. Measures such as the use of face masks (90.4%) and restricted number of patients for access at a time (89.6%) were implemented to reduce the spread of COVID-19. Majority of the respondents (61.6%) believe the measures have reduced the interaction between patients and pharmacists. Most of the respondents (81.6%) believe the pandemic has affected the availability and almost average (48%) of the respondents think that there was inflation in the prices of products like antimalarials, antibiotics, protective and preventive equipment and multivitamins.

**Conclusion:** The COVID-19 pandemic has affected the delivery of pharmaceutical care to clients as well as the availability of medicines to patients. To improve the delivery of pharmaceutical care, tele-pharmacy and online pharmaceutical services may be employed.

## INTRODUCTION

The coronavirus disease 2019 (COVID-19) is a global health pandemic that has resulted in a surge in the demand for frontline healthcare personnel to meet patient care needs. It is a newly identified coronavirus that causes infectious sickness (WHO, 2020a). The virus primarily affects the respiratory system. Fever, dry cough, muscle soreness, exhaustion, anorexia and shortness of breath are the most common symptoms (Ozma *et al.*, 2020). Droplets from the nose or mouth are the primary mechanism of transfer from person to person. According to studies, even asymptomatic patients can spread the infection (WHO, 2020b).

In Nigeria, about 206,279 cases of COVID-19 have been confirmed, with 194,167 patients discharged and 2,723 deaths confirmed as at October 4, 2021 (NCDC, 2021c). Vaccines and therapies are being developed by some scientists and the biomedical research community. Currently, short-term remedies for those that require it have centered on the use of existing antivirals (like remdesivir) that have been licensed for other illnesses off-label. Some drugs are now being tested in clinical trials to determine their efficacy and safety in the treatment of COVID-19 (Patient Safety Network, 2020). Examples of these drugs include antiviral agents like molnupiravir and favipiravir, nitazoxanide, niclosamide, immunomodulators (infliximab, abatacept and cenicriviroc) and corticosteroids (Bergman, 2021).

According to WHO (1994), community pharmacists are the most accessible health care professionals to the general populace. Community pharmacists assist patients by filling prescriptions, dispensing medications, educating and assessing patients for chronic drug renewal, providing minor illness consultations, patient monitoring and examination (WHO, 1994).

Community pharmacists are often the first point of contact with the health system for those with COVID-19-related health concerns or those who seek credible information and advice during this pandemic (Hedima *et al.*, 2020). Community pharmacists are facing a lot of issues as a result of the pandemic in terms of ensuring patient care. These include assistance with infection prevention, supply chain management, stockpile prevention, and the dissemination of evidence-based medical information (Hayden and Parkin, 2020). Both pharmacists and patients are affected by these issues.

A previous study by Hedima *et al.* (2020) was concerned with the role of community pharmacies on the frontline of health service delivery against COVID-19. They ignore the important factor of how the pandemic has affected (and is still affecting) the delivery of services by the community pharmacist to patients. COVID-19 pandemic is placing extraordinary and sustained demands on the health system and providers of essential community services (Emanuel *et al.*, 2020). The pharmacist at the frontline plays a role in providing medications and counseling to the patients, therefore community pharmacies remained open during the early active phase of pandemic and continue to stay open to attend to patients and clients.

The rapid evolution of the COVID-19 pandemic has brought about a series of changes in the community pharmacy sector. In relation to the pandemic, the Center for Disease Control (CDC) published guidelines that highlight key activities of the pharmacist in helping to curb the spread of COVID-19 (CDC, 2020b). These guidelines have caused changes to the delivery of services to patients in the pharmacy. One of such guidelines is the practice of social distancing. Pharmacists adapted their premises to achieve this measure by ensuring the use of face masks, installing barriers and floor markers to instruct waiting patients to remain six feet away from the counter or from other patients or staff. Pharmacists also restricted patient numbers for access and implemented delivery services to patients at home, especially elderly patients who have been advised to stay at home (CDC, 2020b).

Therefore, this study is aimed at assessing the effects of COVID-19 on the delivery of pharmaceutical services by community pharmacists and how pharmacists have adapted to the situation in order to satisfy customers and also curb the spread of COVID-19. The main focus includes the delivery of pharmaceutical care to patients, the availability of drugs and the price of drugs amidst the COVID-19 pandemic.

Pharmaceutical care is based on the responsible provision of drug therapy for the purpose of achieving definite outcomes which improve patient’s quality of life. Little or no study has taken into account the effects of the COVID-19 pandemic on community pharmacy services in the aspects of provision of pharmaceutical care and drug availability and prices, as these factors are crucial to both patients and pharmacists. Therefore, this study will be a useful baseline study for other researchers in this field.

The main objective of this study is to evaluate the effects of the coronavirus 2019 pandemic on community pharmacy activities.

The specific objectives are to:

1. Evaluate the changes caused by COVID-19 on community pharmacies with respect to delivery of pharmaceutical care and availability of medications to patients.
2. Assess how these changes (if any) have affected the delivery of pharmaceutical care to patients.
3. Assess how these changes have affected the availability and price of drugs in the pharmacy.
4. Evaluate how pharmacists have adapted to these changes in order to ensure adequate provision of patient care.

This research focuses on community pharmacies in Lagos state. In light of the ongoing pandemic, the study will employ an electronic format to reach the target population.

## MATERIALS AND METHOD

### STUDY SETTING

The study was carried out in Lagos state, located in the south-western part of Nigeria. It is the most populous city in Nigeria and is the center of commercial activities in the country (Campbell, 2012). The survey was administered to registered community pharmacists via the platforms of Association of Community Pharmacists in Nigeria (ACPN) of various local government areas in Lagos state. The ACPN is the technical arm of the Pharmaceutical Society of Nigeria (PSN) committed to empowering every community pharmacist to embrace best pharmacy practice (PSN, 2019). The study was carried out between February and October, 2021.

### STUDY DESIGN

The study design used for this project was cross sectional study design.

### STUDY POPULATION

The participants involved in the study included licensed pharmacists currently working in a community pharmacy, either as the owner, superintendent pharmacist or as the pharmacist in charge.

### SAMPLE SIZE TECHNIQUES

The study involved community pharmacists with a target sample size of 120 respondents conveniently chosen.

### SAMPLING METHOD

The sampling method used in this study was convenience sampling based on the number of community pharmacists who were willing to participate in the study.

### DATA COLLECTION TOOL

The study was carried out during the pandemic; hence an online questionnaire was used. Data collection tool was created using Google Forms software, an online mobile tool which is used to design customized questionnaires. The questionnaire was structured to cover the main topics:

1. Sociodemographic characteristics including gender, age, position in the pharmacy and number of years in practice.
2. Effect of COVID-19 on the delivery of pharmaceutical care.
3. Effect of COVID-19 on drug availability and prices. A copy of the questionnaire is available in Appendix 1.

### DATA COLLECTION

The online questionnaire was administered to community pharmacists in Lagos state and their responses were the data source. The questionnaire was sent via the ACPN online platforms as well as other available community pharmacists’ WhatsApp group chats to allow willing pharmacists to participate in the study. The survey was sent out between June and August, 2021, during which daily reminders were sent to get as many responses as possible.

### DATA ANALYSIS

Data collected was pre-analyzed using the Google Forms software and was further subjected to descriptive statistical analysis using Microsoft Excel software. Descriptive data were presented in tables, bar charts and pie chart. Frequencies and percentages were also used for analysis of data. Comparisons across demographic characteristics were made using Chi square statistics. A probability of 0.05 or less was considered to be significant. For qualitative questions, thematic analysis of responses was performed using appropriate codes.

### ETHICAL CONSIDERATIONS

For this study, ethical approval was obtained from the Health Research Ethics Committee, Lagos University Teaching Hospital (LUTH), Idi-Araba, Lagos, Nigeria, by a Notice of Exemption. The Health Research Committee assigned number was ADM/DCST/HREC/APP/4167. The pharmacists who participated in filling the questionnaires gave their consent. Utmost confidentiality was ensured during and after data collection.

## RESULTS

Consequently, the questionnaires were distributed to one hundred and fifty (150) community pharmacists and one hundred and twenty-five (125) responses were retrieved. The response rate was 83.33%.

### SOCIODEMOGRAPHIC CHARACTERISTICS

Table 1 shows that from a total of 125 respondents, more than half (56%) were female, and with majority falling within the age range of 25-34 years with the mean age of 32.4 ± 4.7 years. Most of the community pharmacist respondents (67.2%) have been practicing for 0-5 years.

**Table 1.** Sociodemographic characteristics of respondents.

| **SOCIODEMOGRAPHIC CHARACTERISTICS** | **FREQUENCY (n=125)** | **PERCENTAGE (%)** |
| --- | --- | --- |
| **Gender** |  |  |
| Female | 70 | 56.0 |
| Male | 55 | 44.0 |
| **Number of Years in Community Pharmacy Practice (years)** |  |  |
| 0-5 | 84 | 67.2 |
| 6-10 | 12 | 9.6 |
| 11-15 | 11 | 8.8 |
| 16-20 | 5 | 4 |
| 21-25 | 5 | 4 |
| 26-30 | 3 | 2.4 |
| 31-35 | 5 | 4 |
| **Age (years)** |  |  |
| <25 | 42 | 33.6 |
| 25- 34 | 47 | 37.6 |
| 35- 44 | 18 | 14.4 |
| 45-54 | 10 | 8.0 |
| 55-64 | 7 | 5.6 |
| >64 | 1 | 0.8 |
| **Mean age = 32.4 ± 4.7 years** |  |  |

Figure 1 below shows that majority of the respondents were locum pharmacists (34%), while 16.8% of the respondents were full time pharmacists.

**Figure 1.**
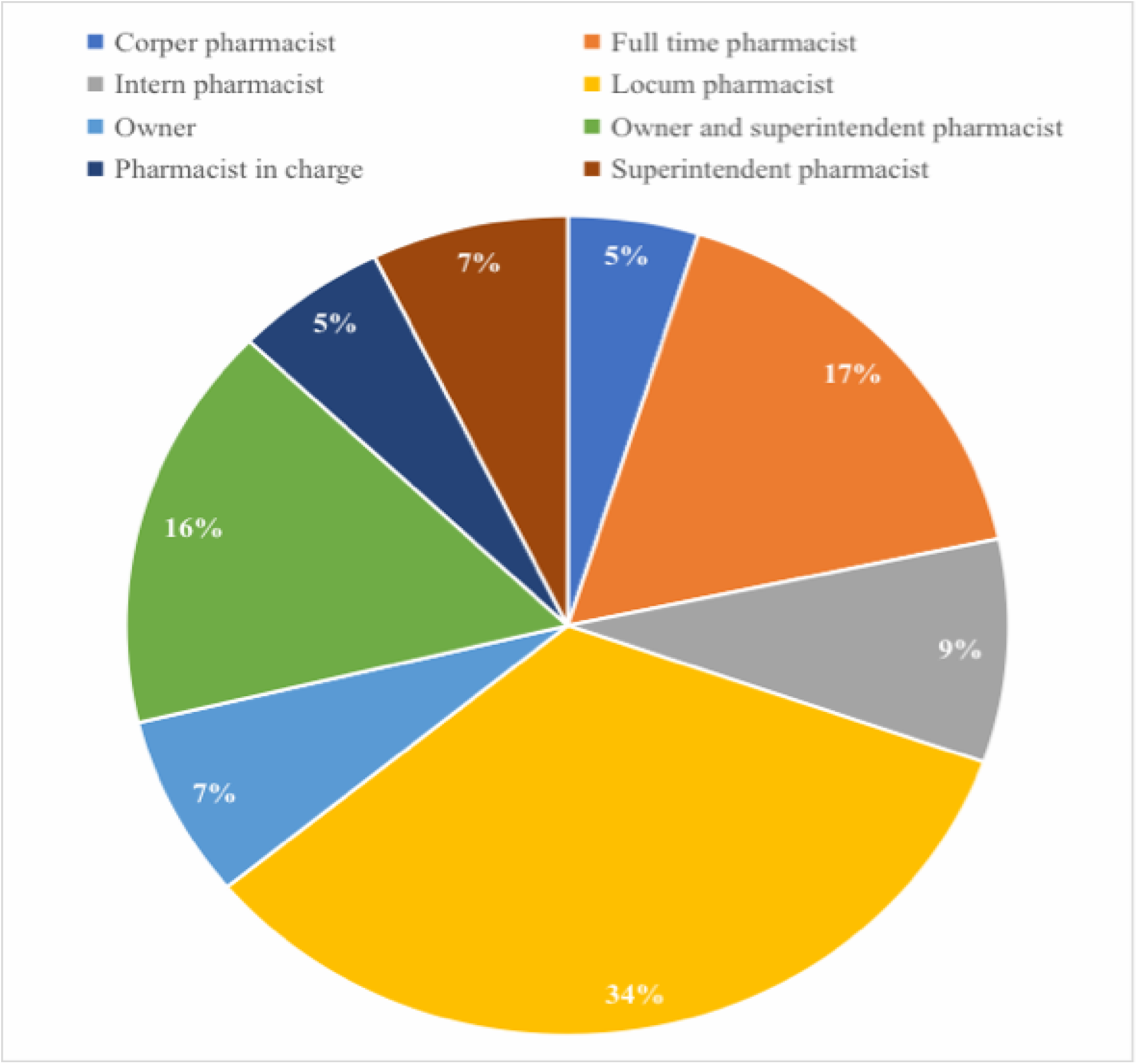
Status of respondents.

### EFFECT OF COVID-19 ON DELIVERY OF PHARMACEUTICAL CARE

The table below shows that the majority of the participants (56%) think there was a total lockdown in the state. Community pharmacies operated during the initial active phase of the pandemic and many (96.8%) of the respondents’ community pharmacies operated during the initial active phase of the pandemic.

**Table 2.**
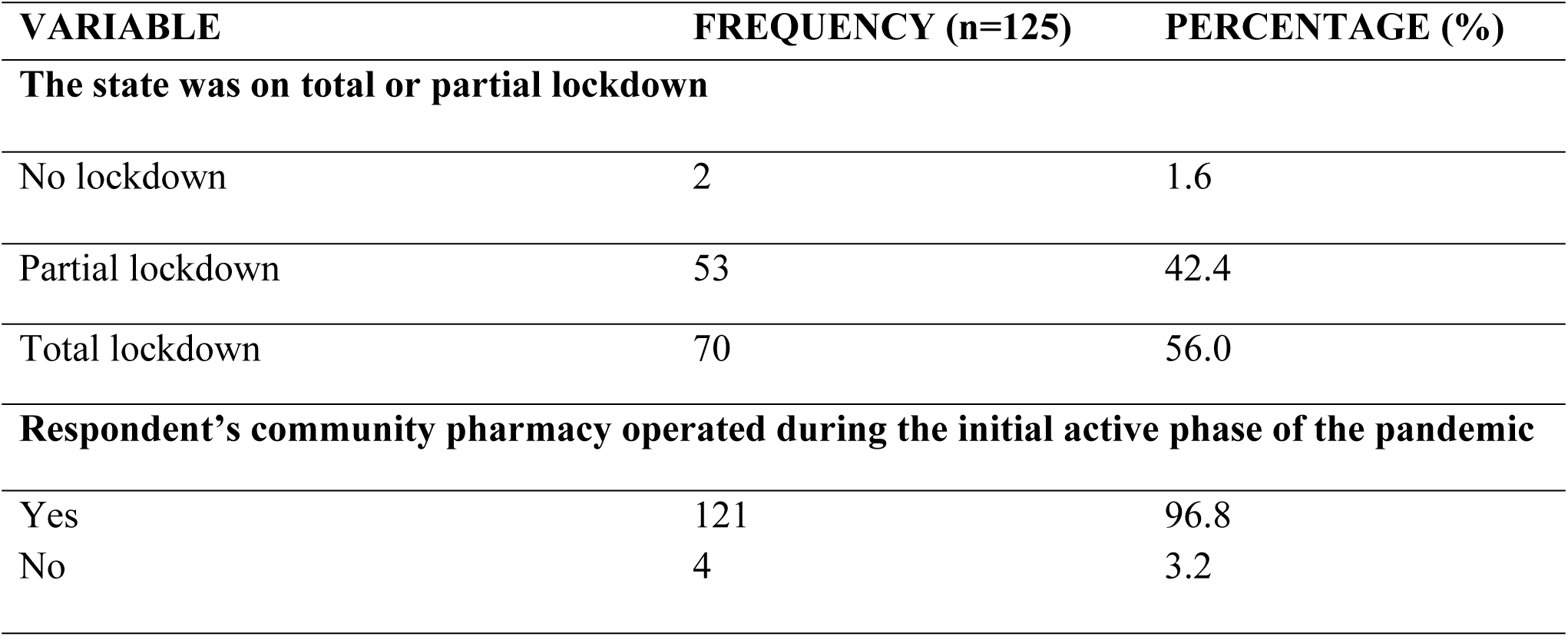
Community pharmacy operation during the lockdown.

Table 3 below shows that most participants practiced the recommended measures to help curb the spread of coronavirus. The most implemented measure was the use of face masks (90.4%). Other commonly practiced measures included restriction in the number of patients for access at a time (90.4%), reduction in number of staff on duty (76.8%) and advice for elderly patients to stay at home (74.4%). Chi square testing against gender and pharmacy status identified that no statistically significant difference exists when compared to the variables.

**Table 3.** Measures adopted during COVID-19.

| **VARIABLE** | **YES**<br>**n (%)** | **NO**<br>**n (%)** | **I DON'T**<br>**KNOW**<br>**n (%)** | **PHARMACY**<br>**STATUS**<br>**(LOCUM**<br>**PHARMACISTS)**<br>**df; p-value** | **GENDER**<br>**(FEMALE);**<br>**df; p-value** |
| --- | --- | --- | --- | --- | --- |
| No face mask, no entry | 113(90.4) | 9(7.2) | 3(2.4) | 1; 0.22 | 1; 0.11 |
| Restricted number of patients for access at a time | 113(90.4) | 9(7.2) | 3(2.4) | 1; 0.06 | 1; 0.44 |
| Reduction in the number of staff on duty | 96(76.8) | 25(20) | 4(3.2) | 1; 0.07 | 1; 0.32 |
| Advice for elderly patients to stay at home | 93(74.4) | 24(19.2) | 8(6.4) | 1; 0.14 | 1; 0.93 |
| Use of markers to ensure 6 feet distance between patients | 76(60.8) | 46(36.8) | 3(2.4) | 1; 0.28 | 1; 0.76 |
| Use of glass barriers to create a distance between patients and the counter | 50(40) | 67(54.4) | 7(5.6) | 1; 0.72 | 1; 0.15 |
| No measures were put in place | 2(1.6) | 114(91.2) | 9(7.2) |  |  |

Table 4 below shows that the measures adopted during the initial active phase of COVID-19 are still being utilized by some pharmacies and many respondents (68.8%) believe the measures against COVID-19 have affected delivery of pharmaceutical care to patients.

**Table 4.** Utilization and effects of measures adopted during the initial active phase of the pandemic.

| **VARIABLE** | **FREQUENCY (n=125)** | **PERCENTAGE (%)** |
| --- | --- | --- |
| **The measures adopted during the pandemic are still being utilized in the operation of your pharmacy** |  |  |
| Yes | 69 | 55.2 |
| No | 56 | 44.8 |
| **The measures adopted during the pandemic affect the quality of patient's care provided in your pharmacy** |  |  |
| Yes | 86 | 68.8 |
| No | 19 | 15.2 |
| I don't know | 20 | 16.0 |

Figure 2 below shows how the measures implemented have affected the delivery of pharmaceutical care to patients. The top three include decreased time spent with patients during counselling (61.6%), pharmacists no longer checking blood pressure and blood sugar level (40%) increased sales of point of care testing devices (38.4%).

**Figure 2.**
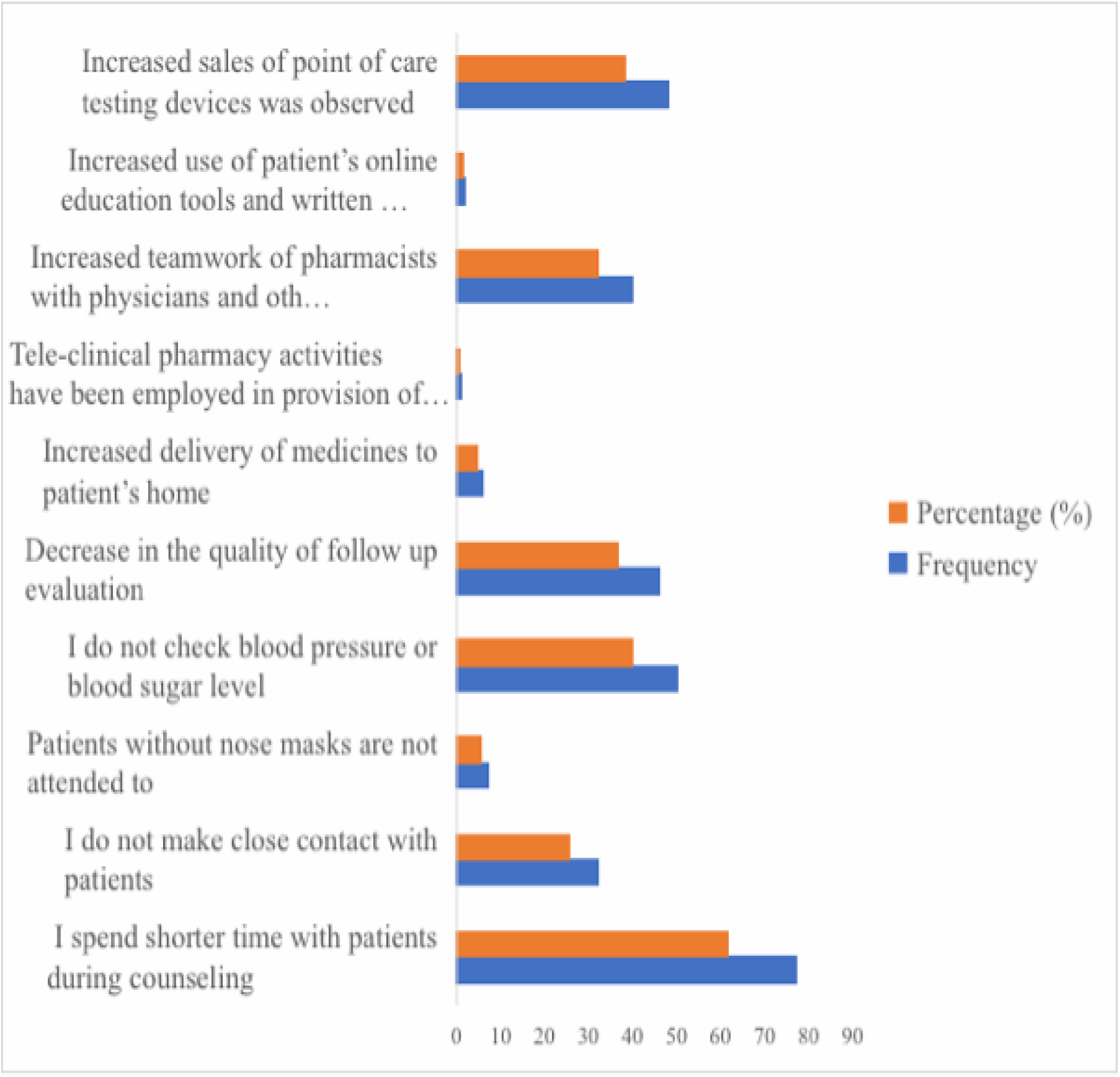
Effect of COVID-19 on the delivery of pharmaceutical care

Table 5 shows that less than half (45.6%) of the respondents believe that the virus has caused moderately positive effects on the pharmaceutical care delivery, while a few of the respondents (32%) believe that there have been moderately negative effects.

**Table 5.** Rate of effect of measures adopted during the pandemic on the patients care delivery.

| **VARIABLE** | **FREQUENCY (n=125)** | **PERCENTAGE (%)** |
| --- | --- | --- |
| **Positive effect of COVID-19 on the delivery of pharmaceutical care by community pharmacists on a scale of 1-5** |  |  |
| 1 (no effect) | 8 | 6.4 |
| 2 (mildly positive effect) | 15 | 12.0 |
| 3 (moderately positive effect) | 57 | 45.6 |
| 4 (positive effect) | 34 | 27.2 |
| 5 (greatly positive effect) | 11 | 8.8 |
| **Negative effect of COVID-19 on the delivery of pharmaceutical care by community pharmacists on a scale of 1-5** |  |  |
| 1 (no effect) | 10 | 8.0 |
| 2 (mildly negative effect) | 18 | 14.4 |
| 3 (moderately negative effect) | 40 | 32.0 |
| 4 (negative effect) | 37 | 29.6 |
| 5 (greatly negative effect) | 20 | 16.0 |

Table 6 shows how respondents have adapted to the changes to ensure delivery of pharmaceutical care. Four thematic areas emerged from respondents on how they ensured the delivery of pharmaceutical care during the pandemic. Many of the respondents ensured the adaptation of the COVID-19 protocols.

**Table 6.** How respondents ensured the delivery of pharmaceutical care during the pandemic.

| **THEMES** | **CODES** | **DEFINITION** |
| --- | --- | --- |

| **The pharmacy adapted to these changes to ensure delivery of pharmaceutical care to patients by** |  |  |
| --- | --- | --- |
| Adapting to the COVID-19 protocols as recommended by the NCDC | ‘Adapting to protocols’<br>‘Implementation of COVID-19 measures’ | Implementation of COVID-19 measures like the use of face masks, hygiene measures and social distancing. |
| Referrals to hospitals | ‘Referrals’ | Referring patients to hospitals for critical cases. |
| Encouraging tele-pharmacy | ‘Tele-pharmacy’<br>‘Calling patients for follow up’<br>‘WhatsApp messages’<br>‘Online’ | The use of WhatsApp messages, calls, videos, and text messages to provide education and information and to follow up on patients. |
| The use of logistics company to offer delivery services for pharmaceutical goods to patients | ‘Home delivery services’<br>‘Logistics’ | Ensuring delivery of medications to clients by the use of logistics especially to elderly patients and ill patients. |
| **THEMES** | **EXPLANATORY NOTES** |  |
| Adapting to the COVID-19 protocols as recommended by the NCDC | Respondents have ensured the implementation of measures of COVID-19 to help limit the spread of the virus. These measures include the use of face masks, hygienic measures and social distancing. |  |
| Referral to hospitals | Respondents have ensured that they refer patients to clinics in critical cases and also to take the COVID-19 test when they feel the symptoms. |  |
| Encouraging tele-pharmacy | Respondents have ensured the use of tele-pharmacy in providing education to patients and in the follow up of patients. |  |
| The use of logistics company to offer delivery services for pharmaceutical goods to patients | The use of logistic services for delivery of medications to elderly patients and ill patients has been implemented by respondents. |  |

### FFECT OF COVID-19 ON DRUG AVAILABILITY AND PRICES

Figure 3 below shows that the COVID-19 has affected the ready availability of medicines in the majority of the respondents’ pharmacies (81.6%).

**Figure 3.**
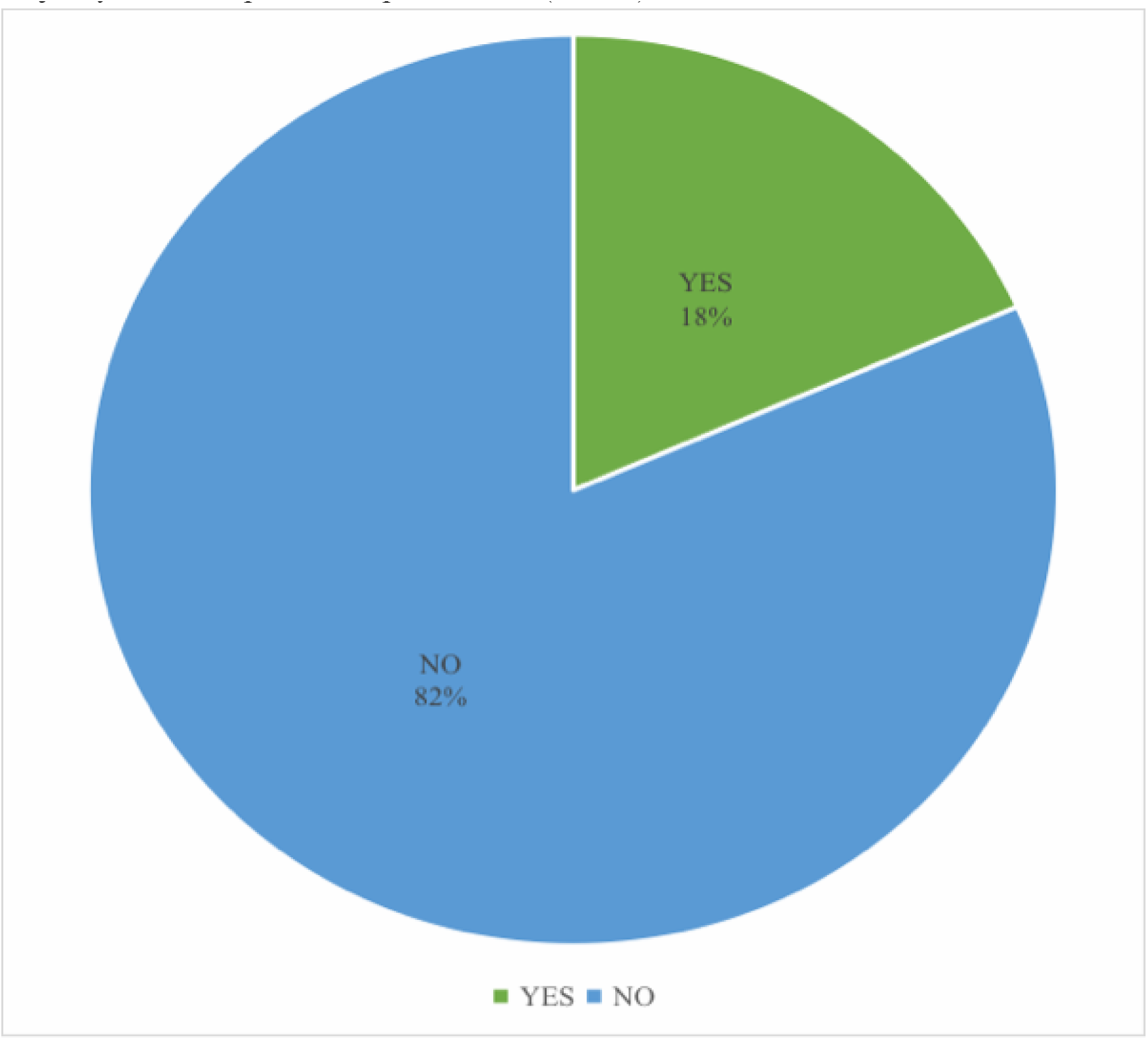
Effect of COVID-19 on the ready availability of medicines in the pharmacy

Table 7 shows that the most affected class of pharmaceutical products was COVID-19 pharmaceutical goods (53.6%). Others were antimalarials (16.8%), antimicrobials (14.4%) and multivitamins (4.7%).

**Table 7.** Classes of medicines most affected.

| **CLASSES OF MEDICINES MOST AFFECTED** | **FREQUENCY (n=125)** | **PERCENTAGE (%)** |
| --- | --- | --- |
| COVID-19 pharmaceutical goods (PPE, hand sanitizers, face masks and immune boosters) | 67 | 53.6 |
| Antimalarials | 21 | 16.8 |
| Antimicrobials | 18 | 14.4 |
| Minerals and Vitamins | 6 | 4.7 |
| Analgesics | 5 | 4.0 |
| Anti-hypertensive | 3 | 2.4 |
| Anti-diabetics | 2 | 1.6 |
| All | 3 | 2.4 |

The medicines that were in short supply in most of the respondents’ pharmacies include antibiotics (24.8%), immune boosters (18.4%) and hydroxychloroquine (12%).

**Table 8.** Medicines Short in Supply.

| **MEDICINES SHORT IN SUPPLY** | **FREQUENCY (n=125)** | **PERCENTAGE (%)** |
| --- | --- | --- |
| Antibiotics | 31 | 24.8 |
| Immune boosters | 23 | 18.4 |
| Hydroxychloroquine | 15 | 12.0 |
| Anti-malarial | 5 | 4.0 |
| Vitamins and Supplements | 2 | 1.6 |
| Anti-hypertensive | 1 | 0.8 |
| Analgesics | 1 | 0.8 |
| Anti-diabetics | 1 | 0.8 |
| Zinc tablets | 1 | 0.8 |
| None | 45 | 36.0 |

Figure 4 shows that some of the respondents (48%) believe that there were dramatic price changes on a few medicines.

**Figure 4.**
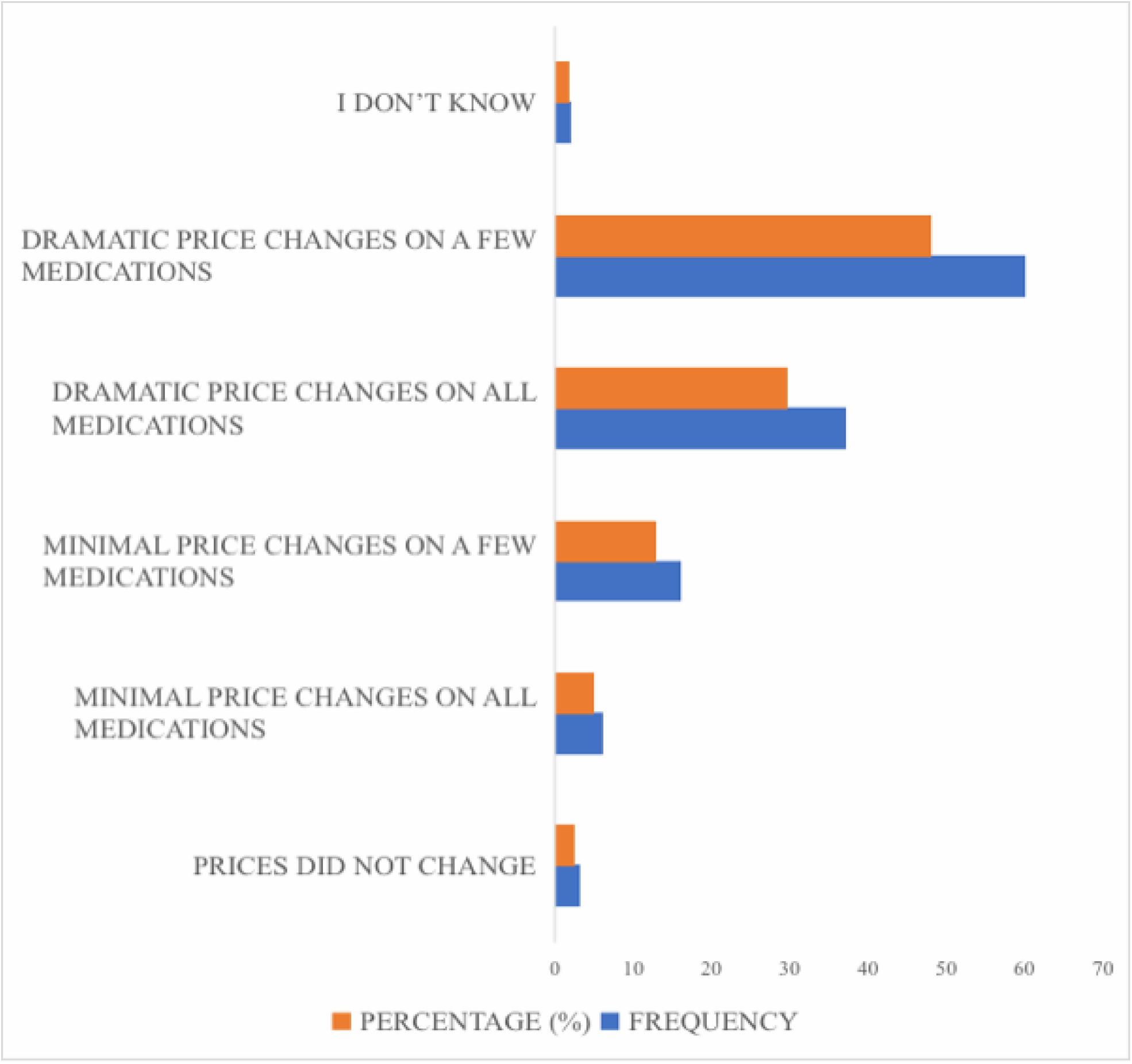
Rating of price changes

Table 9 shows that multivitamins and immune boosters had the most dramatic price changes (28.7%). Other medications with dramatic price changes were antimalarials, antibiotics and ivermectin.

**Table 9.** Medicines with dramatic price changes.

| **MEDICINES WITH DRAMATIC PRICE CHANGES** | **FREQUENCY**<br>**(n=125)** | **PERCENTAGE**<br>**(%)** |
| --- | --- | --- |
| Multivitamins and immune boosters | 53 | 28.7 |
| Antimalarial | 33 | 17.8 |
| Antibiotics | 30 | 16.2 |
| Ivermectin | 27 | 14.6 |
| Preventive or protective equipment | 12 | 6.5 |
| Anti-hypertensive | 8 | 4.3 |
| Antidiabetic | 7 | 3.9 |
| Antipyretics, antihistamines | 6 | 3.2 |
| Analgesic | 2 | 1.1 |
| Infusions and astringent | 2 | 1.1 |
| Antiviral | 1 | 0.5 |
| Antifungal | 1 | 0.5 |
| Asthma drugs | 1 | 0.5 |
| Imported drugs | 1 | 0.5 |
| Almost all | 1 | 0.5 |
| None | 2 | 1.1 |

From Table 10 below, the majority of the respondents (18.4%) were constantly able to contact distributors for supply of drugs during the initial active phase of the pandemic.

**Table 10.** Ways by which respondents were able to meet patients’ drug supply needs during the initial active phase of the pandemic.

| **HOW PATIENTS' DRUG SUPPLY NEEDS WERE MET** | **FREQUENCY**<br>**(N=125)** | **PERCENTAGE**<br>**(%)** |
| --- | --- | --- |
| Ask from distributors | 23 | 18.4 |
| We had enough stock | 17 | 13.6 |
| Through phone contacts and recommendations | 12 | 9.6 |
| Bulk purchasing | 18 | 14.4 |
| Delivery services were introduced and clients paid on delivery | 6 | 4.8 |
| Better alliance with suppliers | 1 | 0.8 |
| Tele-pharmacy | 1 | 0.8 |
| I don't know | 47 | 37.6 |

## DISCUSSION

This study was carried out with the aim of assessing the effect of COVID-19 on community pharmacy service delivery. Of 125 participants, 56% were female. This is similar to a study in the Netherlands which had 64.7% female participants (Koster *et al.*, 2020b). The modal age range was 25-34 years with a mean age of 32.4 ± 4.7 years. This was in contrast with the study in the Netherlands which had a mean age of 43.4 ± 11.5 years (Koster *et al.*, 2020b). A great percentage of the respondents were locum pharmacists and only few of the respondents were pharmacy owners. This is in contrast to a study carried out in Australia where the majority of the participants were full-time pharmacists and only a few were locum pharmacists (Sum and Ow, 2020). This could be due to the use of online platforms containing more locum pharmacists and less pharmacy owners. Majority of the respondents have had 0-5 years of practice. This is similar to a study carried out in Switzerland in which 79% of the respondents have had <5 years of experience as community pharmacists (Hoti *et al.*, 2020).

The lockdown in Lagos state took effect on March 30, 2020 (Mbah, 2020). Majority of the respondents’ community pharmacies were actively operating during this period. This shows that during the pandemic, community pharmacists were the first point of contact for individuals and they remained accessible to patients, providing health related information and advice (Sum and Ow, 2020).

Actions in response to coronavirus were mainly aimed at reducing direct patient contact and limiting the number of patients visiting the pharmacy. The findings show that the measures implemented by the NCDC and CDC were implemented in most community pharmacies. However, there was a variability in the frequency of uptake of some of these measures. For example, the use of face masks, restriction of the number of patients for access at a time, reduction in number of staff on duty and advice for elderly patients to stay at home were implemented by the majority of the pharmacies. The use of glass barriers, however, was less frequently implemented. The study carried out in the Netherlands showed that community pharmacists suggested the use of plastic or glass barriers as a standard for hygiene in the pharmacy (Koster *et al.*, 2020b). Majority of the community pharmacists ensured the use of face masks in the pharmacy. The use of face masks has been proved to be effective in reducing transmission of the coronavirus (Esposito *et al.*, 2020). More than half of the respondents indicated that the measures are still in use.

The study also reveals that more than half of the pharmacists believed the pandemic has affected the delivery of pharmaceutical care to patients. A good number of the pharmacists believe that the effects have been moderately positive (45.6%) and moderately negative (32%). Majority (61.6%) of the community pharmacists now spend shorter time with patients during counseling. This is in contrast to the study carried out in Australia, where only a few pharmacists found the provision of clinical services and medication counseling challenging (Sum and Ow, 2020). The structural increase in the distance between pharmacists and patients negatively influences patient-pharmacist interaction (Koster *et al.*, 2020b). Some respondents have had to stop checking blood pressure and blood sugar level. This could also be due to physical distancing. Decrease in the quality of follow up evaluation has also been noticed. This could be due to the inadequate patient-pharmacist time, physical distancing and advice for elderly patients and ill to stay home. Social and physical distancing may compromise the quality of patient care and medication counseling provided in community pharmacists. However, to ensure the safety of patients and staff, the use of tele-pharmacy has been encouraged (Sum and Ow, 2020). Tele-pharmacy and use of online education tools have been promoted to facilitate communication between pharmacists and their patients (Liu *et al.*, 2020). However, only very few of the respondents were able to employ the use of tele-pharmacy and online pharmacy. The methods are difficult because many patients have limited digital skills as well as health literacy and therefore are at risk of drug related problems (Koster *et al.*, 2020a).

Also, the use of logistic procedures to deliver drugs to patients have been implemented. This is essential especially for high risk patients such as elderly patients and critically ill patients (Koster *et al.*, 2020b). Overall, many of the community pharmacists ensured the practice of the COVID-19 protocols recommended by the NCDC.

Majority of the respondents agreed that COVID-19 has affected the ready availability of medicines in their pharmacies. This could be as a result of limited importation of goods from other countries (mainly China and India) and the panic buying practice of customers in response to the pandemic (Uwizeyimana *et al.*, 2021). Also the price increase from pharmaceutical wholesalers and patients buying unnecessary and excessive products have also contributed to the unavailability of some drugs (Hoti *et al.*, 2020). The COVID-19 pharmaceutical goods were the most affected class of medicines including face masks, sanitizers and preventive or protective equipment (PPE). This is similar to a study conducted in Bangladesh, where over 95% of the stores recorded an increase in the purchase of PPE (Haque *et al.*, 2020). Other medicines like antimalarials and antimicrobials were also affected. The antibiotics and immune boosters were short in supply in most pharmacies due to increased demand for them. This is similar to a study carried out in developing countries, where it was shown that there was an increase in utilization of antibiotics and multivitamins in Nigeria, Pakistan, Bangladesh, Ghana and Vietnam (Sefah *et al.*, 2021). Antimalarials, principally hydroxychloroquine, were also short in supply in some pharmacies (12%). This is also similar to the study carried out in developing countries, where there was an increase in utilization of hydroxychloroquine across most African countries except Namibia (Sefah *et al.*, 2021). This is as a result of its use as a prophylaxis or treatment of COVID-19 despite the controversies against its usefulness (WHO, 2020h).

Findings from this study shows that there were dramatic price changes on a few medicines, especially the immune boosters and antimalarial medicines. This is similar to the study conducted in Bangladesh where dramatic price changes were also noticed. This is as a result of the decrease in importation and increase in utilization during the pandemic (Haque *et al.*, 2020). Increase in local production could help address the situation (EAC, 2020).

## CONCLUSION

The research findings reveal that the coronavirus disease 2019 (COVID-19) had considerable impact on the delivery of pharmaceutical care to patients and on the availability of drugs in community pharmacies in Lagos state. Although the lockdown was quite challenging, community pharmacy continued to provide services by implementing measures to reduce the spread of COVID-19. These measures caused a decrease in interaction between pharmacists and their patients. To improve delivery of patient care, pharmacists identified the importance of logistics services and tele-pharmacy in healthcare. Provision of drugs and other pharmaceutical products was challenging, but pharmacists were able to ensure steady delivery of drugs by ensuring bulk purchasing and always contacting suppliers.

## Data Availability

All data produced in the present work are contained in the manuscript

## Notes

### Competing Interest Statement

The authors have declared no competing interest.

### Funding Statement

This study did not receive any funding

### Author Declarations

Ethical approval was obtained from Health Research Ethics Committee, Lagos University Teaching Hospital (LUTH), Idi-Araba, Lagos State

### Summary of Updates

This version of the manuscript has been revised to update the author list and the author affiliations.

